# HIV prevention and missed opportunities among people with recently acquired HIV infection: Α protocol for a systematic review

**DOI:** 10.1101/2023.12.05.23299513

**Authors:** Argyro Karakosta, Elisa Ruiz-Burga, Shema Tariq, Giota Touloumi, Emily Jay Nicholls, Nikos Pantazis, Inma Jarrin, Marc Van der Valk, Caroline Sabin, Christina Mussini, Laurence Meyer, Alain Volny Anne, Christina Carlander, Sophie Grabar, Linda Wittkop, Bruno Spire, Jonh Gill, Kholoud Porter, Fiona Burns, CASCADE Collaboration

## Abstract

**Background:** Individuals who have recently acquired HIV represent a unique population because the time frame since HIV acquisition is relatively short and identification of missed HIV prevention opportunities is, therefore, closer to real time and less subject to recall bias. Identifying prevention measures used and missed opportunities for using them, can help stop further HIV transmission.

**Objectives:** This systematic review aims to synthesise current global evidence on uptake of HIV prevention methods among people with recently acquired HIV from 2007, the year that the concept of ART as a prevention method was first introduced.

**Methods and analysis:** MEDLINE/PubMed, EMBASE, PsycINFO, Cochrane and Web of Science databases, will be searched for articles published January 2007 - July 2023. Eligible studies will be those that reported on HIV prevention methods among people with recently acquired HIV. Quality assessment of the studies selected will be undertaken, and reporting of the systematic review will be informed by Preferred Reporting Items for Systematic Reviews and Meta-Analyses (PRISMA) guidelines.

**Results:** The results of the systematic review will be available by the end of January 2024.

**Conclusions:** The findings will be of key relevance to researchers, healthcare providers including third sector organisations/ community groups and policymakers, as they will offer insight into better understanding of missed or failed HIV prevention efforts and will help ensure future efforts meet the needs of those in need of them.

## Introduction

HIV prevention has changed dramatically over the past three decades and effective combination prevention, that includes Pre-Exposure Prophylaxis (PrEP) and treatment as prevention (TasP) [1,2] as components, means elimination of HIV transmission is now feasible. As a consequence, prevention is a key component of the ambitious agenda set in 2021 by the global HIV community, “End Inequalities. End AIDS”. As part of the United Nations Declaration on Ending AIDS [3], Member States also committed to 95% of people at risk of HIV infection having access to appropriate, person-centred and effective combination prevention options by 2025. However, despite antiretroviral therapy (ART) being freely available to people living with HIV for many years [1] and widening access to PrEP [4,5], HIV transmission remains high, with around 1.5 million people estimated to have acquired HIV in 2021 [6]. Multiple HIV prevention strategies exist to reduce the risk of HIV acquisition and transmission; however, they remain underutilised [7].

Acute HIV infection (AHI, defined here as the first four weeks following HIV acquisition), and primary HIV infection (PHI, defined here as the six-month period following acquisition) [8], are a period of high infectiousness and may contribute disproportionately to ongoing HIV transmission [9]. People with recently acquired HIV are most likely to accurately recall the circumstances around acquisition, allowing identification and understanding of missed opportunities for HIV prevention. Studies of individuals with recently acquired HIV can provide useful insights for HIV prevention efforts and lead to a decline of transmission.

We propose undertaking a systematic review to synthesise current global evidence on prior access to and uptake of HIV prevention interventions among people with recently acquired HIV. To prevent duplication of reviews, a preliminary search of similar protocols and reviews was conducted in July 2023 using the Cochrane Library, MEDLINE, Embase, PubMed, Web of Science, PsycINFO and The International Prospective Register of Systematic Reviews (PROSPERO) databases. No review protocol or systematic review on this topic was identified. Rather, systematic reviews on recently acquired HIV, identified through searches of these databases, examined the diagnostic, clinical and public health implications of identifying and treating people with recently acquired HIV [10-18]. A consistent theme among these studies was the important contribution of people with recently acquired HIV to HIV epidemics, as several groups have reported disproportionate rates of onward HIV transmission from individuals with acute infection, although this issue remains controversial [11]. According to these studies, transmission clusters tend to be driven by recently acquired undiagnosed infection, and although estimates are highly variable, recently acquired HIV has been identified as the source for between 10% and 50% of all transmissions [18]. During the earliest stages of HIV infection plasma viral load levels increase exponentially, and this comprises a dominant factor predicting transmission to sexual partners [19]. In addition, discrepancies in the reported contribution of recently acquired HIV infection to ongoing epidemics could be explained by differences in epidemic stage, definitions of “early HIV”, and variation in sexual behaviours (i.e., anal vs. vaginal intercourse, partner change rates, etc.) [13,18,19]. Even in a well-established HIV epidemic where the role of recently acquired HIV generally appears to be lower, detection of persons with recently acquired HIV provides an opportunity to intervene at the earliest possible stage of infection [12] to reduce the risk of missed opportunities for prevention of onward HIV transmission.

A better understanding of missed or failed HIV prevention opportunities will help ensure prevention efforts better meet those in need of them. This systematic review aims to synthesise current global evidence on uptake of HIV prevention methods among people with recently acquired HIV. Specifically, this systematic review seeks to answer the following questions: 1) What HIV prevention methods have people with recently acquired HIV used, if any? 2) What are the structural and behavioural barriers to the uptake, use of, and adherence to HIV prevention methods?

## Materials and Methods

This will be a systematic review designed to summarise evidence of prior access to and uptake of HIV prevention methods among people who had recently acquired HIV infection. The review will commence in September 2023. This protocol has been developed in accordance with the Preferred Reporting Items for Systematic Review and Meta-Analysis Protocols (PRISMA-P) guidelines [20] [Appendix 1] and reporting of the synthesised findings will also be informed by PRISMA guidelines [21]. Additionally, this protocol is registered in PROSPERO (CRD42023454414). Important amendments to this protocol will be published along with the results of the systematic review.

For this review, recently acquired HIV will mean HIV acquisition within the 12 months preceding HIV diagnosis. For each included study we will document the criteria used to establish recency, with the gold standard being the availability of an HIV-negative antibody test within 12 months of the first positive one, or other laboratory evidence of acute HIV infection.

### Searches

We will search peer reviewed articles on the access and use of HIV prevention interventions among people with recent HIV infection, published in English, from 1 January 2007 to 17 July 2023 (2007 being the year that the concept of ART as a prevention method was introduced [22]). From our search, we will identify studies that assessed at least one of: HIV prevention, condom use, uptake of HIV testing, PrEP, post exposure prophylaxis (PEP), harm reduction (i.e., needle exchange, opioid substitution therapy (OSP), etc.), counselling. The search will be performed in the following databases:

- CINAHL Plus
- MEDLINE
- PubMed
- PsycINFO
- Web of Science
- EMBASE

The following search strategy will be adapted for all databases.

Searches will combine key terms relating to HIV/AIDS (HIV, AIDS, human immunodeficiency virus, etc.) with terms related to recently acquired HIV (seroconverter, acute infection, recent infection, recently acquired, etc.) and terms related to preventions methods (condoms, preexposure prophylaxis, HIV testing, counselling, treatment, combination prevention, etc.). See Appendix 2 for full search strategy.

### Types of study to be included

All types of epidemiological studies including randomised controlled trials (RCTs), case-control studies, and cohort studies, both with and without a control group, will be included. RCTs where the intervention is some type of prevention will not be considered, except if baseline information about prevention is mentioned. Qualitative studies will be excluded as our group has recently published a systematic review of qualitative literature on recently acquired HIV [8]. Studies that are not peer-reviewed will also be excluded. Studies employing mixed methods approaches will be included where the quantitative component is presented in sufficient detail. We will not include grey literature or review articles.

### Participants/population

This review will include studies from any country on adults aged 16 years and over who have recently acquired HIV and where use of at least one HIV prevention method is measured.

The review will exclude studies which focus on people who have recently been diagnosed with HIV, but where the time of HIV acquisition is not known, or if no HIV prevention method is mentioned. We will exclude studies in those aged under 16.

### Intervention(s), exposure(s)

The focus of this review is on prevention methods among people with recently acquired HIV. Preliminary engagement with the literature suggests that this will include: mapping of current HIV prevention methods among people with recently acquired HIV and factors affecting the use of prevention measures (address barriers to uptake, use of, and adherence to strategies to prevent HIV transmission).

### Main outcome(s)

The expected outcomes are identifying the prevention methods used, and not used, by people with recently acquired HIV and identifying the barriers to uptake of, use of, and adherence to strategies to prevent HIV acquisition.

### Eligibility criteria

This review will include studies which report on adults aged 16 years and over who have recently acquired HIV and where use of at least one HIV prevention method is measured, published between 1/1/2007 and 31/7/2023, and written in English.

### Exclusion criteria

No known time of HIV acquisition/ seroconversion, younger than 16 years of age, published earlier than 2007 or recruitment of population earlier than 2007. Studies combining adults and paediatric/adolescent patients will be included only if able to disaggregate results by age to identify those aged ≥16.

### Study selection

Potential articles will be collected in the systematic review software Covidence [23]. Duplicates will be removed by AK using Covidence software. Based on the pre-specified eligibility criteria, studies will be selected using a two-staged approach; titles and abstracts will be examined first and then the full-text of all potential eligible studies will be retrieved and screened. AK and ER-B will independently screen a sample of 100 citations to pre-test and refine coding guidance based on the inclusion criteria and until at least 90% agreement is achieved. Disagreements about eligibility will be resolved through discussion. AK will then screen the remaining citations for inclusion in the review using the pre-tested coding guidance. Full-text of all potentially eligible studies will be retrieved and AK and ER-B will assess all full text articles to determine final study selection. Differences will be resolved through consensus, with input on eligibility from a third reviewer (FB) when necessary. Citations that do not meet the inclusion criteria will be excluded and the reason for exclusion will be recorded at the full-text screening.

### Assessment of risk of bias and quality of evidence

The quality of each included study will be assessed by using the Newcastle Ottawa Scale (NOS) for cohort studies and modified NOS for cross-sectional and case-control studies [24]. We assume that there will be no or very few RCTs on this topic and we will, therefore, use the National Institute for Health and Care Excellence (UK) Quality Appraisal of Intervention Studies tool (derived from Jackson et al., 2006 [25]) as a risk of bias assessment tool. Each paper included in the review will be quality assessed by AK and ER-B. All disagreements will be resolved through consensus. FB will be consulted to discuss any discrepancies in study quality assessment.

### Data extraction

Data will be extracted from the final articles using a standardised form in Covidence. This will include: 1) characteristics of the study: author, year of publication, study objective, country and context, 2) methodological characteristics: study design, research questions and/or hypotheses, study population, definition of recently acquired HIV used, sample characteristics, type of prevention’/s’ measure/s mentioned, 3) main findings and conclusions.

### Data synthesis

A narrative (descriptive) synthesis of the data will be conducted, after a final list of papers has been compiled. This discussion will be tailored around the type of prevention strategy, target population characteristics, barriers to uptake, use of and adherence to prevention strategies.

Provided the studies are sufficiently homogeneous and comparable regarding study design, percentages will be pooled through meta-analysis or even meta-regression to explain part of the variability.

Reporting of the systematic review will be informed by Preferred Reporting Items for Systematic Reviews and Meta-Analyses (PRISMA) guidelines [21], as mentioned before.

### PROSPERO registration number

CRD42023454414

### Patient and Public Involvement (PPI)

Community members have been involved in the conception and design of this protocol and will contribute to interpretation and dissemination of findings.

### Ethics and dissemination

Ethics approval is not required for a systematic review. Findings of the systematic review will be disseminated through open access publication in a peer-reviewed journal. The findings will be of interest to researchers, healthcare practitioners, policymakers, and HIV community partners.

## Results

This review will be undertaken as part of the CASCADE Collaboration, a mixed methods international study investigating the medical and lived experiences of people with recently acquired HIV (https://www.cascadestudy.net/) [26]. We expect that the review will be completed by the end of January 2024. We will report the results based on the identified outcomes as specified above.

## Discussion

Despite new and effective interventions and declines in HIV incidence across Western Europe, a significant number of people continue to acquire HIV. In this systematic review, we are committed to producing evidence that addresses barriers in access to HIV prevention. People who have recently acquired HIV are a unique, and ideal group, in whom to study these clinically and public health relevant questions, and to assess prevention methods used. We believe that our data are extremely relevant to ongoing HIV prevention efforts.

The review will provide evidence to inform the provision of HIV treatment and prevention services. The expected outcomes are identifying the prevention methods used, whether they are used appropriately and effectively by people with recently acquired HIV, and examining the structural and behavioural barriers to uptake, use of, and adherence to HIV prevention measures.

## Data Availability

No datasets were generated or analysed during the current study.

## Authors contribution

The study protocol was conceptualised by FB, KP, AK and ER-B contributed to protocol design and development of the rationale and methodology. AK wrote the first draft of the manuscript. FB and KP provided critical feedback on overall study design and manuscript. All authors read, revised and approved the final manuscript.

## Collaborators

CASCADE Collaboration: CASCADE Executive Committee: Santiago Moreno (Chair), Fiona Burns, Rafael Eduardo Campo, Harmony Garges, Cristina Mussini, Nikos Pantazis, Barbara Pinto, Kholoud Porter, Caroline Sabin, Shema Tariq, Giota Touloumi, Vani Vannappagari, Alain Volny Anne, Lital Young. CASCADE Scientific Steering Committee: John Gill (co-chair), Kholoud Porter (co-chair), Christina Carlander, Rafael Eduardo Campo, Harmony Garges, Sophie Grabar, Inma Jarrín, Laurence Meyer, Barbara Pinto, Giota Touloumi, Marc van der Valk, Vani Vannappagari, Alain Volny Anne, Linda Wittkop, Lital Young. CASCADE Social Science subcommittee: Shema Tariq (Chair), Agnes Aisam, Diana Barger, Udi Davidovich, Marie Dos Santos, Lars Eriksson, Eli Fitzgerald, John Gill, Sophie Grabar, Inma Jarrín, Argyro Karakosta, Hartmut Krentz, Cristina Mussini, Emily Jay Nicholls, Nicoletta Policek, Elisa Ruiz-Burga, Chris Sandford, Bruno Spire, Inés Suárez-García, Giota Touloumi, Alain Volny Anne.

## Supporting Information

S1 Appendix 1. PRISMA-P (preferred reporting items for systematic review and meta-analysis protocols) 2015 checklist: recommended items to address in a systematic review protocol.

S2 Appendix 2. Full systematic review database search string

## Abbreviations

HIV: Human Immunodeficiency Virus
ART: Antiretroviral Therapy
PrEP: Pre-Exposure Prophylaxis
TasP: Treatment as Prevention
AHI: Acute HIV Infection
PHI: Primary HIV Infection
PRISMA-P: Preferred Reporting Items for Systematic Review and Meta-Analysis Protocols
PRISMA: Preferred Reporting Items for Systematic Review and Meta-Analysis
PEP: Post Exposure Prophylaxis
OSP: Opioid Substitution Therapy
RCTs: Randomized controlled trials
NOS: Newcastle Ottawa Scale

## Supporting Information

### S1 Appendix 1

**PRISMA-P (preferred reporting items for systematic review and meta-analysis protocols) 2015 checklist: recommended items to address in a systematic review protocol**.

| Section and Topic | Item # | Checklist item | Location where the item is reported (#page) |
| --- | --- | --- | --- |
| **ADMINISTRATIVE INFORMATION** |  |  |  |
| **Title:** |  |  |  |
| Identification | 1a | Identify the report as a protocol of a systematic review | 1 |
| Update | 1b | If the protocol is for an update of a previous systematic review, identify as such | NA |
| **Registration** | 2 | If registered, provide the name of the registry (such as PROSPERO) and registration number | 5,7 |
| **Authors:** |  |  |  |
| Contact | 3a | Provide name, institutional affiliation, e-mail address of all protocol authors; provide physical mailing address of corresponding author | 1 |
| Contributions | 3b | Describe contributions of protocol authors and identify the guarantor of the review | 8 |
| **Amendments** | 4 | If the protocol represents an amendment of a previously completed or published protocol, identify as such and list changes; otherwise, state plan for documenting important protocol amendments | NA |
| **Support:** |  |  |  |
| Sources | 5a | Indicate sources of financial or other support for the review | 2 |
| Sponsor | 5b | Provide name for the review funder and/or sponsor | 2 |
| Role of sponsor of funder | 5c | Describe roles of funder(s), sponsor(s), and/or institution(s), if any, in developing the protocol | 2 |
| **INTRODUCTION** |  |  |  |
| **Rational** | 6 | Describe the rationale for the review in the context of what is already known | 4,5 |
| **Objectives** | 7 | Provide an explicit statement of the question(s) the review will reference to participants, interventions, comparators, and outcomes (PICO) | 6 |

| METHODS |  |  |  |
| --- | --- | --- | --- |
| **Eligibility criteria** | 8 | Specify the study characteristics (such as PICO, study design, setting, time frame) and report characteristics (such as years considered, language, publication status) to be used as criteria for eligibility for the review | 5,6 |
| **Information sources** | 9 | Describe all intended information sources (such as electronic databases, contact with study authors, trial registers or other grey literature sources) with planned dates of coverage. | 5,6 |
| **Search strategy** | 10 | Present draft of search strategy to be used for at least one electronic database, including planned limits, such that it could be repeated | 5, Appendix 2 |
| **Study records:** |  |  |  |
| Data management | 11a | Describe the mechanism(s) that will be used to manage records and data throughout the review | 6,7 |
| Selection process | 11b | State the process that will be used for selecting studies (such as two independent reviewers) through each phase of the review (that is, screening, eligibility and inclusion in meta-analysis) | 6,7 |
| Data collection process | 11c | Describe planned method of extracting data from reports (such as piloting forms, done independently, in duplicate), any processes for obtaining and confirming data from investigators | 7 |
| **Data items** | 12 | List and define all variables for which data will be sought (such as PICO items, funding sources), any pre-planned data assumptions and simplifications. | 7 |
| **Outcomes and prioritization** | 13 | List and define all outcomes for which data will be sought, including prioritization of main and additional outcomes, with rationale | 6 |
| **Risk of bias in individual studies** | 14 | Describe anticipated methods for assessing risk of bias of individual studies, including whether this will be done at the outcome or study level, or both; state how this information will be used in data synthesis. | 6,7 |
| **Data synthesis** | 15a | Describe criteria under which study data will be quantitatively synthesised | 7 |
| | 15b | If data are appropriate for quantitative synthesis, describe planned summary measures, methods of handling data and methods of combining data from studies, including any planned exploration of consistency (such as $I^2$, Kendall's T) | 7 |
|  | 15c | Describe any proposed additional analyses (such as sensitivity or subgroup analyses, meta-regression) | NA |
|  | 15d | If quantitative synthesis is not appropriate, describe the type of summary planned | 7 |
| **Meta-bias(es)** | 16 | Specify any planned assessment of meta-bias(es) (such as publication bias across studies, selective reporting within studies) | NA |
| **Confidence in cumulative evidence** | 17 | Describe how the strength of the body of evidence will be assessed (such as GRADE) | 6,7 |

*From:* <u>Shamseer</u> L, Moher D, Clarke M, Ghersi D, Liberati A, Petticrew M, Shekelle P, Stewart LA, the PRISMA-P Group. Preferred reporting items for systematic review and meta-analysis protocols (PRISMA-P) 2015: elaboration and explanation. BMJ. 2015;349:g7647.

### S2 Appendix 2. Full systematic review database search string

**Table A.** Search strings for Ovid Medline (in-process & other non-indexed citations and Ovid MEDLINE)

| # | Search string |
| --- | --- |
| 1 | HIV infections/ or HIV seropositivity/ |
| 2 | HIV.ab,kw,ti. |
| 3 | "Human Immunodeficiency Virus".ab,kw,ti. |
| 4 | 1 or 2 or 3 |
| 5 | ((recent* or new*) adj3 (infect* or acqui*)).ab,kw,ti. |
| 6 | (seroconver* or sero-conver* or recent* HIV infect* or acute infect* or acute HIV or primary HIV).ab,kw,ti. |
| 7 | 5 or 6 |
| 8 | (prevent* or PREP or pre-exposure or preexposure or PEP or post exposure or condom* or harm reduction or risk reduction or needle* exchange or opioid substitut* or syringe or HIV test* or behaviour or behavior or counselling or counseling or HIV transmission or missed opportunit* or treatment or antiretroviral* or ART or sex* or comb* prevent* or TasP).ab,kw,ti. |
| 9 | 4 and 7 and 8 |
| 10 | limit 9 to (yr="2007 -Current" and "humans only (removes records about animals)") |

**Table B.**
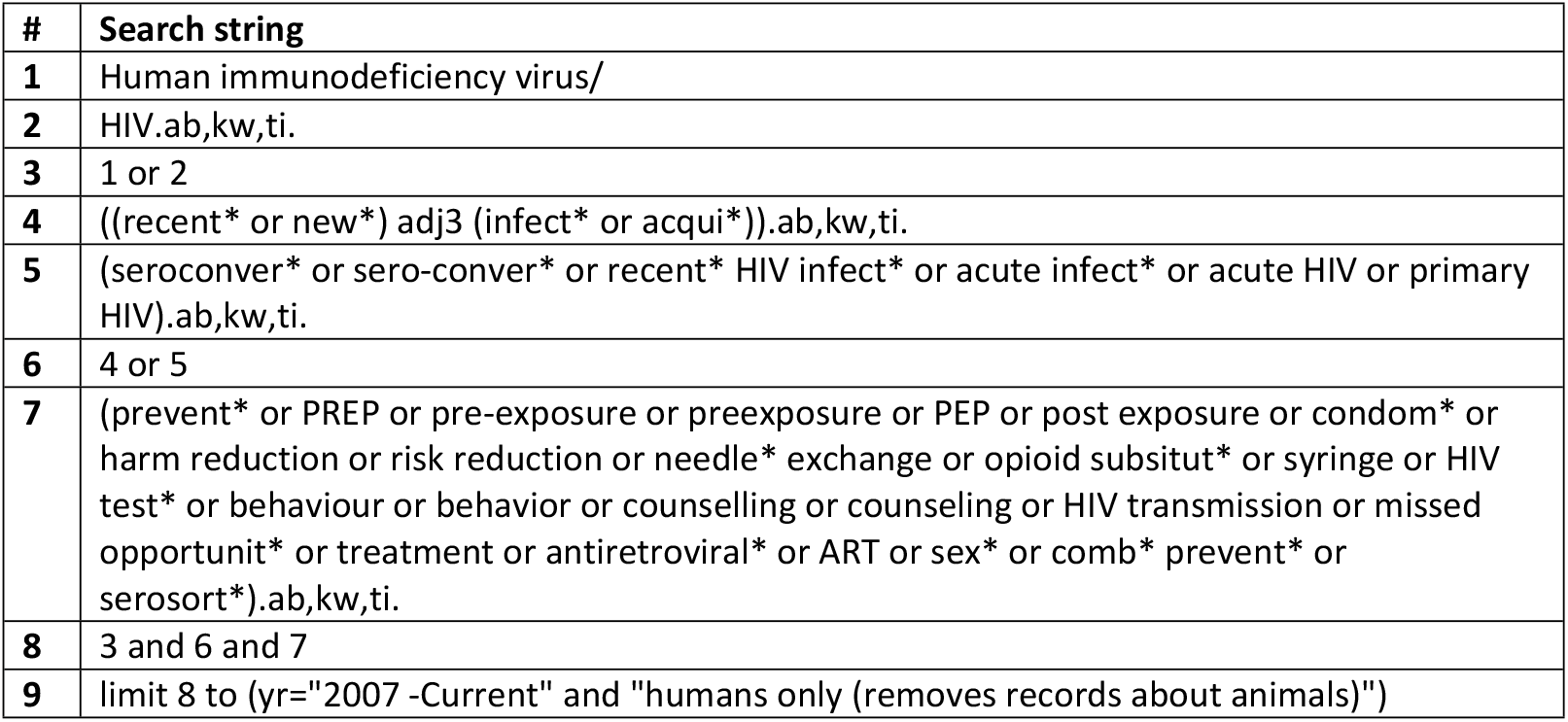
Search strings for Embase.

**Table C.** Search strings for PubMed.

| # | Search string |
| --- | --- |
| 1 | HIV[MeSH Terms] |
| 2 | HIV seropositivity[MeSH Terms] |
| 3 | human immunodeficiency virus[Title/Abstract] |
| 4 | HIV infections[MeSH Terms] |
| 5 | (#1 OR #2 OR #3 OR #4) |
| 6 | recent* infect*[Title/Abstract] OR recent* acqui*[Title/Abstract] OR new* infect*[Title/Abstract] OR new* acqui*[Title/Abstract] OR acute infect*[Title/Abstract] OR |
|  | seroconver*[Title/Abstract] OR sero-conver*[Title/Abstract] OR recent* HIV infect*[Title/Abstract] OR acute HIV[Title/Abstract] OR primary HIV[Title/Abstract] |
| **7** | prevent*[Title/Abstract] OR PREP[Title/Abstract] OR pre-exposure[Title/Abstract] OR preexposure[Title/Abstract] OR PEP[Title/Abstract] OR post exposure[Title/Abstract] OR condom*[Title/Abstract] OR harm reduction[Title/Abstract] OR risk reduction[Title/Abstract] OR needle* exchange[Title/Abstract] OR opioid substitut*[Title/Abstract] OR syringe[Title/Abstract] OR HIV test*[Title/Abstract] OR behaviour[Title/Abstract] OR behavior[Title/Abstract] OR counselling[Title/Abstract] OR counseling[Title/Abstract] OR HIV transmission[Title/Abstract] OR missed opportunit*[Title/Abstract] OR treatment[Title/Abstract] OR antiretroviral*[Title/Abstract] OR ART[Title/Abstract] OR sex*[Title/Abstract] OR comb* prevent*[Title/Abstract] OR TasP[Title/Abstract] |
| **8** | (#5 AND # 6 AND #7) |
| **9** | Humans, Publication date from 2007/1/1 |

**Table D.**
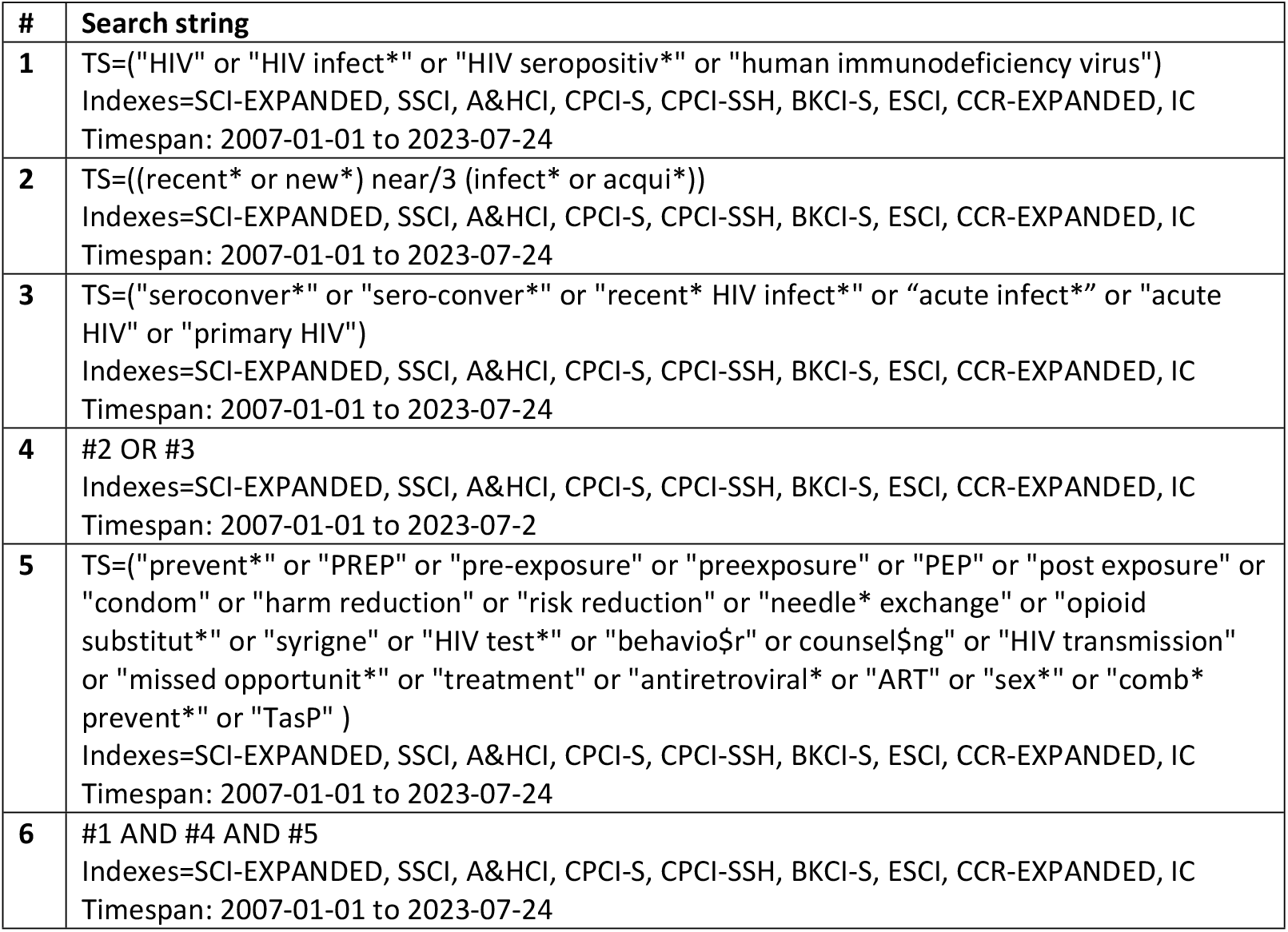
Search strings for Web of Science.

**Table E.** Search strings for the Cochrane Library.

| # | Search string |
| --- | --- |
| **1** | MeSH descriptor: [HIV] explode all trees |
| **2** | MeSH descriptor: [HIV Seropositivity] explode all trees |
| **3** | "HIV" OR HIV NEXT infect* OR HIV NEXT seporosit* OR "human immunodeficiency" (Word variations have been searched) |
| **4** | MeSH descriptor: [Seroconversion] explode all trees |
| 5 | (recent* NEXT infect* OR recent* AND acqui* OR new* AND infect* OR new* NEXT acqui* OR acute NEXT infect* OR seroconver* OR sero-conver* OR recent* NEXT HIV NEXT infect* OR "acute HIV" OR "primary HIV"):ti,ab,kw (Word variations have been searched) |
| 6 | MeSH descriptor: [Pre-Exposure Prophylaxis] explode all trees |
| 7 | MeSH descriptor: [Post-Exposure Prophylaxis] explode all trees |
| 8 | MeSH descriptor: [Condoms] explode all trees |
| 9 | MeSH descriptor: [Harm Reduction] explode all trees |
| 10 | MeSH descriptor: [Risk Reduction Behavior] explode all trees |
| 11 | MeSH descriptor: [Needle-Exchange Programs] explode all trees |
| 12 | MeSH descriptor: [Opiate Substitution Treatment] explode all trees |
| 13 | MeSH descriptor: [HIV Testing] explode all trees |
| 14 | MeSH descriptor: [Anti-Retroviral Agents] explode all trees |
| 15 | (prevent* OR PREP OR pre-exposure OR preexposure OR PEP OR "post exposure" OR condom* OR "harm reduction" OR "risk reduction" OR needle* NEXT exchange OR opioid NEXT substitut* OR syringe OR HIV NEXT test* OR behaviour OR behavior OR counselling OR counseling OR "HIV transmission" OR missed NEXT opportunit* OR treatment OR antiretroviral* OR ART OR sex* OR comb* NEXT prevent* OR TasP):ti,ab,kw |
| 16 | #1 OR #2 OR #3 |
| 17 | #4 OR #5 |
| 18 | #6 OR #7 OR #8 OR #9 OR #10 OR #11 OR #12 OR #13 OR #14 OR #15 |
| 19 | #17 AND #18 AND #19 (with Cochrane Library publication date from Jan 2007 to Aug 2023, in Cochrane Reviews, Cochrane Protocols, Trials, Clinical Answers, Editorials and Special Collections (Word variations have been searched) |

**Table F.** Search strings for PsycINFO.

| # | Search string |
| --- | --- |
| 1 | exp HIV/ |
| 2 | ((recent* or new*) adj3 (infect* or acqui*)).ab,ti. |
| 3 | (seroconver* or sero-conver* or recent* HIV infect* or acute infect* or acute HIV or primary HIV).ab,ti. |
| 4 | 2 or 3 |
| 5 | (prevent* or PREP or pre-exposure or preexposure or PEP or post exposure or condom* or harm reduction or risk reduction or needle* exchange or opioid substitut* or syringe or HIV test* or behaviour or behavior or counselling or counseling or HIV transmission or missed opportunit* or treatment or antiretroviral* or ART or sex* or comb* prevent* or TasP).ab,ti. |
| 6 | 1 and 4 and 5 |
| 7 | limit 6 to yr="2007 -Current" |

## Notes

**Conflicts of interest** The funders did not participate in the study design and will not intervene in its process, analysis or publication of the findings. ST has received speaker honoraria and consultancy fees from Gilead Sciences. CC has received speaker/moderator honoraria and advisory board fees from Gilead Sciences, GSK/ViiV and MSD as well as an unrestricted Gilead Sciences Nordic Fellowship Research Grant. CS has received funding from Gilead Sciences, ViiV Healthcare and Janssen-Cilag for participation in Advisory Boards, speaker panels and for preparation of educational materials. MVdV has received consultancies fees for participation in advisory boards and research grants from Gilead, MSD and ViiV all paid to his institution. FB has received funding from Gilead Sciences Ltd for preparation and delivery of educational materials. IJ has received teaching fees from ViiV Healthcare and advisory fees from Gilead Sciences. GT has received research grants and advisory board fees from Gilead, all paid to her institution, and MJG has received honoraria for ad hoc participation in national Advisory boards of Gilead Merck and ViiV.

### Competing Interest Statement

Conflicts of interest The funders did not participate in the study design and will not intervene in its process, analysis or publication of the findings. ST has received speaker honoraria and consultancy fees from Gilead Sciences. CC has received speaker/moderator honoraria and advisory board fees from Gilead Sciences, GSK/ViiV and MSD as well as an unrestricted Gilead Sciences Nordic Fellowship Research Grant. CS has received funding from Gilead Sciences, ViiV Healthcare and Janssen-Cilag for participation in Advisory Boards, speaker panels and for preparation of educational materials. MVdV has received consultancies fees for participation in advisory boards and research grants from Gilead, MSD and ViiV all paid to his institution. FB has received funding from Gilead Sciences Ltd for preparation and delivery of educational materials. IJ has received teaching fees from ViiV Healthcare and advisory fees from Gilead Sciences. GT has received research grants and advisory board fees from Gilead, all paid to her institution, and MJG has received honoraria for ad hoc participation in national Advisory boards of Gilead Merck and ViiV.

### Funding Statement

Yes

### Author Declarations

Ethics approval is not required for a systematic review.

